# Effects of Topical Anesthetics on catheter-related bladder Discomfort in patients undergoing ureteroscopic litholapaxy: A Single-Center Randomized Controlled Study

**DOI:** 10.64898/2026.04.04.26350148

**Authors:** Chengzhong Ma, Mengxiao Wei, Zuheng Wang, Xuemei Li, Yanling Feng, Yuee Luo, Xiao Lu, Weiru Wang, Shijia Zhou, Xiuping Li, Fubo Wang, Wenwu Liu

**Affiliations:** Department of Urology, Guangxi International Zhuang Medicine Hospital, Nanning, Guangxi, China; Department of Tuberculosis, The Fourth People’s Hospital of Nanning, Nanning, Guangxi, China; Department of Urology, the First Affiliated Hospital of Guangxi Medical University, Guangxi Medical University, Nanning, Guangxi, China; Department of Rrproductive Health and Infertility, Guangxi International Zhuang Medicine Hospital Affiliated to Guangxi niversity of Chinese Medicine, Nanning, Guangxi, China; Department of Medicine, Faculty of Chinese Medicine Science, Guangxi University of Chinese Medicine, Nanning, Guangxi, China

**Keywords:** Topical anesthesia, Ureteroscopic lithotripsy, Catheter-related bladder discomfort, urethra pain NRS scores

## Abstract

**Background:** Urinary catheterization is a routine procedure after ureteroscopy lithotripsy (URSL), but it often causes catheter-related bladder discomfort (CRBD) and urethral pain, which aggravates patients’ postoperative discomfort. This study finds out the effect of topical anesthesia on CRBD and urethra pain in patients undergoing ureteroscopy lithotripsy and urinary catheterization.

**Methods:** In this study, 330 patients undergoing ureteroscopy lithotripsy enrolled, with 160 cases in the control group and 170 cases in the experimental group. The experimental group divided into two subgroups based on the local anesthetic used: Tetracaine Hydrochloride Gel subgroup and Oxybuprocaine Gel subgroup. Postoperative assessments conducted using CRBD scores and urethra pain numerical rating scale (NRS) score. CRBD and urethra pain NRS scores measured at T0, T1, T2, T3, T4, T5, and T6.

**Results:** Compared to the control group, the use of local anesthetics significantly reduced both CRBD scores and urethra pain NRS scores in the experimental group, with the differences being statistically significant (P < 0.01). In male patients, patients who used local anesthetics markedly decreased CRBD scores and urethra pain NRS scores compared to those not receiving local anesthetics, showing statistical significance (P < 0.01), whereas no significant difference followed in female patients. No statistically significant differences found between Rigid ureteroscopy lithotripsy (R-URSL) and Flexible ureteroscopy lithotripsy (F-URSL) regardless of the use of local anesthetics. Within the experimental group, the effects of different local anesthetics were similar, with comparable impacts on CRBD scores and urethra pain NRS scores, and no statistical differences noted. These findings suggest that local anesthetics are effective in reducing postoperative CRBD scores and urethra pain NRS scores, especially in male patients.

**Conclusion:** Topical anesthesia following ureteroscopy lithotripsy reduces CRBD scores and urethra pain NRS scores in patients undergoing urinary catheterization, especially in male patients.

**Trial registration:** Chinese Clinical Trial Registry (No. ChiCTR2500105477, https://www.chictr.org.cn/. date: July 4, 2025).

## Background

Catheter-related bladder discomfort (CRBD) is a common clinical issue faced following urinary catheterization, with an incidence rate ranging from 47% to 90% [1–3]. This condition especially pronounced in men, especially after transurethral surgery [4–7], with 27% to 55% classified as moderate to severe [2]. CRBD can charge to various reasons, including the irritative effects of the catheter on the bladder mucosa, forming of biofilms, and the inflammatory response triggered by foreign bodies. Other reasons include catheter size, catheter material, stone fragments, patient age, and so on [5, 8–11].These reasons contribute to discomfort and may lead to increased urinary urgency, frequency, and pain. CRBD characterized as a burning sensation, radiating from the suprapubic region to the penis, and associated with discomfort or a strong urge to void. Its symptoms closely resemble those of an overactive bladder (OAB) [1, 2, 5, 12]. Additionally, CRBD often associate to behavioral responses such as flailing limbs, vocal expression of distress, and tries to remove the urinary catheter [1]. These clinical shows of CRBD may affect the recovery and satisfaction during treatment [1]. Mitigating CRBD is therefore a critical for improving patient care and satisfaction. Effective management strategies are essential not only for relieving symptoms but also for promoting faster recovery and improving overall patient results.

CRBD arises from catheter-induced irritation of the bladder. To reduce these symptoms, several pharmacological interventions have explored. Muscarinic receptor antagonists, analgesic, sedative and local anesthetics, such as butylscopolamine, tolterodine, tramadol, nefopam, Gabapentin, dexmedetomidine, lidocaine and Oxybutynin, have proved efficacy in reducing CRBD symptom [1, 2, 5, 7, 12]. Our study focused on two local anesthetic agents, Tetracaine Hydrochloride Gel and Oxybuprocaine Gel. Tetracaine is an ester-type local anesthetic, metabolized in the plasma and liver by plasma pseudocholinesterase [13]. Tetracaine commnnly used in medical settings for spinal anesthesia, as well as a local anesthetic for diagnostic purposes and examinations [13, 14]. When applied topically, it is 5 to 8 times more potent than cocaine[13]. Oxybuprocaine, an ester of para-amino-benzoic acid, widely used in ophthalmology as a topical anesthetic [15]. The 0.4% Oxybuprocaine hydrochloride ophthalmic solution commonly used in Europe for topical corneal anesthesia in both large and small animals, with effectiveness evaluated in dogs, cats, horses, and cattle [15].

Ureteroscopy lithotripsy (URSL), both rigid and flexible, is a common procedure for treating of upper urinary tract calculi. However, due to transurethral and the frequent postoperative placement of ureteral stents and catheterization, patients undergoing URSL are at an increased risk of CRBD and urethra pain [10, 11, 16]. Despite its prevalence, there is a notable lack of research focusing on CRBD and urethra pain following URSL. This gap highlights the innovative of our study, which mainly explores the role of local anesthetics in mitigating CRBD and urethra pain after ureteroscopy lithotripsy. The findings may contribute to relieve of CRBD symptoms and urethra pain in patients undergoing ureteroscopy lithotripsy.

## Methods

### Study design

This study is a Randomized Controlled Trial conducted at Guangxi International Zhuang Medicine Hospital. All patients enrolled between September 20^th^, 2024 to July 18^th^, 2025. This research has been approved by the Medical Ethics Committee of Guangxi International Zhuang Medical Hospital.

Inclusion criteria for this study were as follows: (1) a confirmed diagnosis of upper urinary tract stones (ureteral or renal stones) based on clinical and radiological assessments. (2) underwent ureteroscopy lithotripsy (rigid or flexible); (3) aged between 18 and 85 years. (4) an American Society of Anesthesiologists (ASA) physical status classification of 1 or 2. (5) Informed consent for participation.

Exclusion criteria encompassed patients with an ASA classification of 3 or higher, those who received conservative management, extracorporeal shock wave lithotripsy (ESWL), or percutaneous nephrolithotomy (PCNL); and individuals with OAB, liver dysfunction or drug allergies. This study reviewed patient records to extract relevant data on treatment results and associated complications to explain the efficacy and safety of the procedure within a real-world clinical setting.

### Patient randomization

This was a single-center, randomized, double-blind, placebo-controlled, parallel-group trial. We randomly allocate in sequence by number. Eligible participants were randomized in a 1:1 ratio to receive either the control group or the experimental group. The experimental group were randomized in a 1:1 ratio to receive either the Tetracaine Hydrochloride Gel subgroup or Oxybuprocaine Gel subgroup. Given the nature of the interventions, blinding of participants and treating physicians was not feasible; however, outcome assessors and data analysts were blinded to treatment allocation to minimize detection bias.

### Catheterization

All postoperative patients in this study uniformly equipped with F16 silicone urinary catheters, sourced from Zhejiang Medical Devices Co., Ltd. and received F6 ureteral stents of Bard. Catheterization procedures needed that all patients have their bladders emptied immediately after operation. In the control group, before catheterization, paraffin oil fully lubricated the catheter. In contrast, the experimental group received a different approach: Tetracaine Hydrochloride Gel (3g per tube), produced by Zhen’ao Jinyinhua Pharmaceutical Co., Ltd. (National Drug Approval Code: H20123292), and Oxybuprocaine Gel (30mg per tube), manufactured by Shenyang Oasis Pharmaceutical Co., Ltd. (National Drug Approval Code: H21023203), injected into the urethra. Each individual dose consisted of 3g and 30mg, respectively. After a 2-minute interval, catheterization performed. Before catheterization, paraffin oil and Topical Anesthetics(Tetracaine Hydrochloride Gel –3g, Oxybuprocaine Gel-30mg) fully lubricated the catheter. Catheters in both groups inflated with a 10 mL water balloon and kept for 6 to 72 hours, with most of patients having catheters in place for 8-12 hours. This design ensured uniformity in catheter materials and procedures while allowing to evaluate of different methods to ruduce CRBD and urethra pain NRS scores.

Patients who underwent URSL under general anesthesia had an F16 Foley catheter and an F6 ureteral stent placed. Ureteral balloon dilation not routinely required for ureteroscopic procedures. Further, to ensure consistency, all flexible ureteroscopies performed with a standardized F12 common sheath (manufacturer: [Hangzhou Zhexin Medical Devices Co., Ltd], model: [12Fr*450 or 12Fr*350]).

### Assessments

In this study, the primary result measure was the CRBD score, with the secondary result measure of the urethra pain NRS scores. The CRBD score evaluate based on a specific set of criteria: patients experiencing no discomfort assigned 0 points; those with slight discomfort, which was only noticeable when prompted, given 1 point; those experiencing moderate discomfort, characterized by symptoms such as frequent urination, urgency, and lower abdominal distension that were difficult to endure, received 2 points; patients with severe discomfort, involving intolerable distension, severe urethral pain, frequent urination with significant restlessness, and the need for catheter removal, assigned 3 points. The urethra pain NRS scores assessed using a different scale: no urethral pain scores 0 piont; mild pain-Scores ranged from 1 to 3; moderate pain from 4 to 6; and severe pain from 7 to 10.

As well as the primary and secondary measures, a comprehensive set of added data collected, including signs such as age, weight, ASA grade, operation duration, anesthesia duration, Operation type, heart rate (HR), respiratory rate (RR), systolic blood pressure (SBP), diastolic blood pressure (DBP), pulse pressure (PP), mean arterial pressure (MAP) and oxygen saturation (SpO_2_). Patient’s anthropometrics such as height and weight also recorded. And, information on the presence of hypertension and diabetes documented to provide a thorough context of each patient’s health status.

Data collection meticulously scheduled at standardized time points to ensure consistency and reliability. Measurements taken at the following intervals about the patients regaining consciousness post-anesthesia: extubation (T0); 0.5 hours (T1); 1.5 hours (T2); 2.5 hours (T3); 3.5 hours (T4); 4.5 hours (T5); and 5.5 hours (T6) after awakening. All recordings conducted following the removal of the endotracheal tube to ensure the data reflect the patient’s condition in a post-anesthesia state.

### Statistical analysis

All data analyzed using SPSS Statistics software (version 26.0) and R software (version 4.4.2). The figures created using GraphPad Prism 9.0. Quantitative data assessed for distribution. Data conforming to a normal distribution represented by the mean and standard deviation, and the Student’s t-test used for analysis. Data not conforming to a normal distribution represented by the median and interquartile range (IQR), and the Mann-Whitney U test used for statistical analysis. Qualitative data presented as figures and percentages and analyzed using the chi-square test. A bilateral P value < 0.05 considered statistically significan.

## Results

### 1. Baseline of patients

A total of 403 cases were included in this study. After exclusion, 35 cases were eliminated. Among them, 21 cases did not meeting inclusion criteria (URSL could not be performed and changed to another surgical method or treatment approach), 11 cases Declined to participate, and 3 cases were Other reasons (Due to time conflicts and other reasons, the surgery was refused). The remaining 368 cases were randomly assigned to the experimental group(184 cases) and the control group(184 cases). In the control group, 24 cases did not receive allocated intervention after operation, included 12 cases of high fever(T≥38.5°C, Among them, there were 3 cases of Urosepsis), 9 cases of gastrointestinal discomfort in, 3 cases of abnormal heart and lung functions, and 2 cases of other reasons. The experimental group divided in Hydrochloride Gel subgroup(92 cases) and Oxybuprocaine Gel subgroup(92 cases). 10 cases(7 cases of high fever(T≥38.5°C, included 2 cases of Urosepsis), 2 cases of gastrointestinal discomfort, 1 cases of abnormal heart and lung functions) in Hydrochloride Gel subgroup and 4 cases (2 cases of high fever(T≥38.5°C), 1 cases of gastrointestinal discomfort, 1 cases of abnormal heart and lung functions)in Oxybuprocaine Gel subgroup were excluded from this study. Ultimately, this study collected a total of 330 cases, with 160 cases in the control group and 170 cases in the experimental group(82 cases in Hydrochloride Gel subgroup and 88 cases in Oxybuprocaine Gel subgroup)(Fig. 1).

**Fig. 1.**
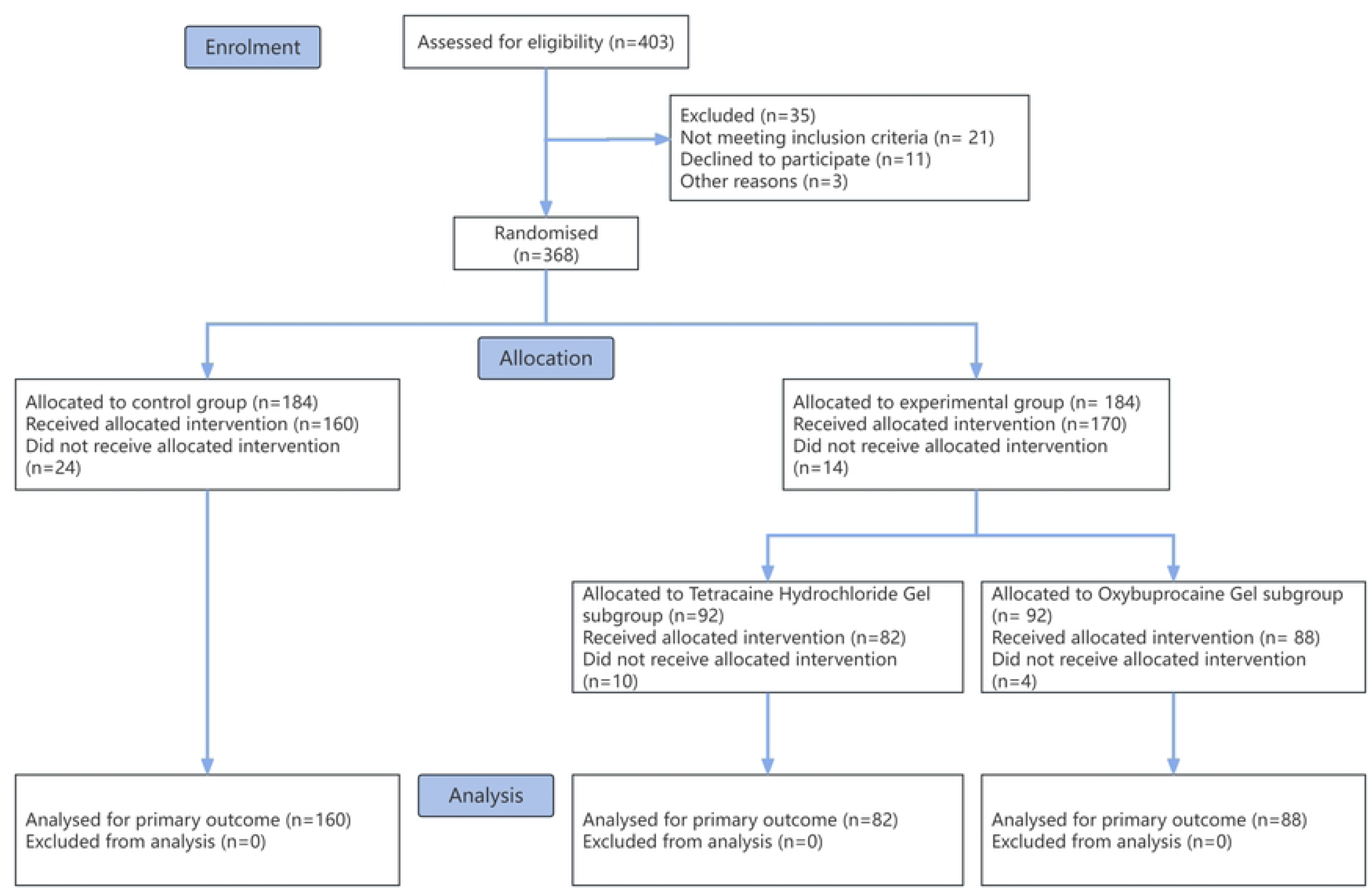
Flowchart of the study.

Out of a total of 330 cases, the age range of the patients spanned from 17 to 84 years, with a mean age of 49 years. The body weights of the patients varied from a minimum of 39 kg to a maximum of 102 kg, with an average weight of 66.8 kg. The duration of anesthesia ranged from 21 to 374 minutes, with a mean of 100 minutes. The surgical duration varied from 7 to 239 minutes, with an average of 57.5 minutes. The time from the end of surgery to full recovery ranged from 0 to 69 minutes, with a mean time of 6.5 minutes. The clinical characteristics were comparable between the control and experimental groups, and consistent within the subgroups of the experimental group. There was no significant difference between the experimental and control groups with age, weight, surgical approach, anesthesia type, diabetes, and hypertension (P > 0.05, table 1). Male patients made up a higher proportion in the experimental group (P < 0.05). Additionally, there was no significant difference in SBP and PP during most of the postoperative period (P > 0.05); however, DBP and MAP was significantly higher in the experimental group for most of the time (P < 0.05).

**Table 1.** Descriptive variables of the control group and experimental group.

### 2. Local anesthetics reduced CRBD and NRS scores

In the present study, primary (CRBD scores) and secondary (NRS score) results measured in both groups. Compared to the control group, the experimental group displayed a statistically significant reduction in CRBD scores from T0 to T6 (all P < 0.01, Fig. 2A). This finding suggests the intervention managed to the experimental group was effective in significantly decline CRBD scores about the control group. The most notable decrease in CRBD scores occurred at the T2-T5 stage, showing the intervention had a helpful pronounced effect at this time point. Consistent with the CRBD score findings described above, compared to the control group, NRS scores were significantly lower in the experimental group from T0 to T6 (all P < 0.01, Fig. 2B), with the most noted difference observed during the T1-T3 phase.

**Fig. 2.**
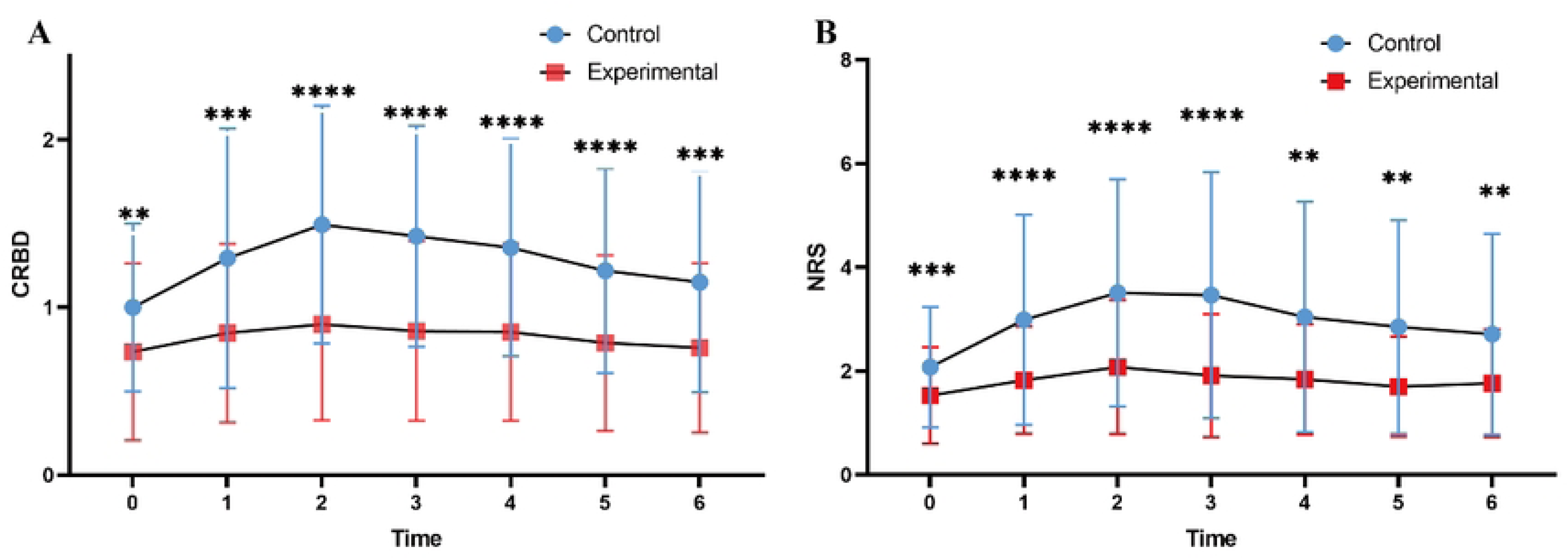
Local anesthetics reduced CRBD (A) and NRS (B) scores. CRBD, catheter-related bladder discomfort; NRS, numerical rating scale.

### 3. Anesthetics were effective for male patients, but not for female

We conducted further analysis by classifying specific subgroups. The study revealed the use of topical anesthetics significantly reduced both CRBD scores and NRS scores in the male subgroup from T0 to T6 (all P < 0.001, Fig. 3A, B). However, in the female subgroup, administrating of anesthetics did not lead to a statistically significant decline in these scores for most of the time (Fig. 3C, D).

**Fig. 3.**
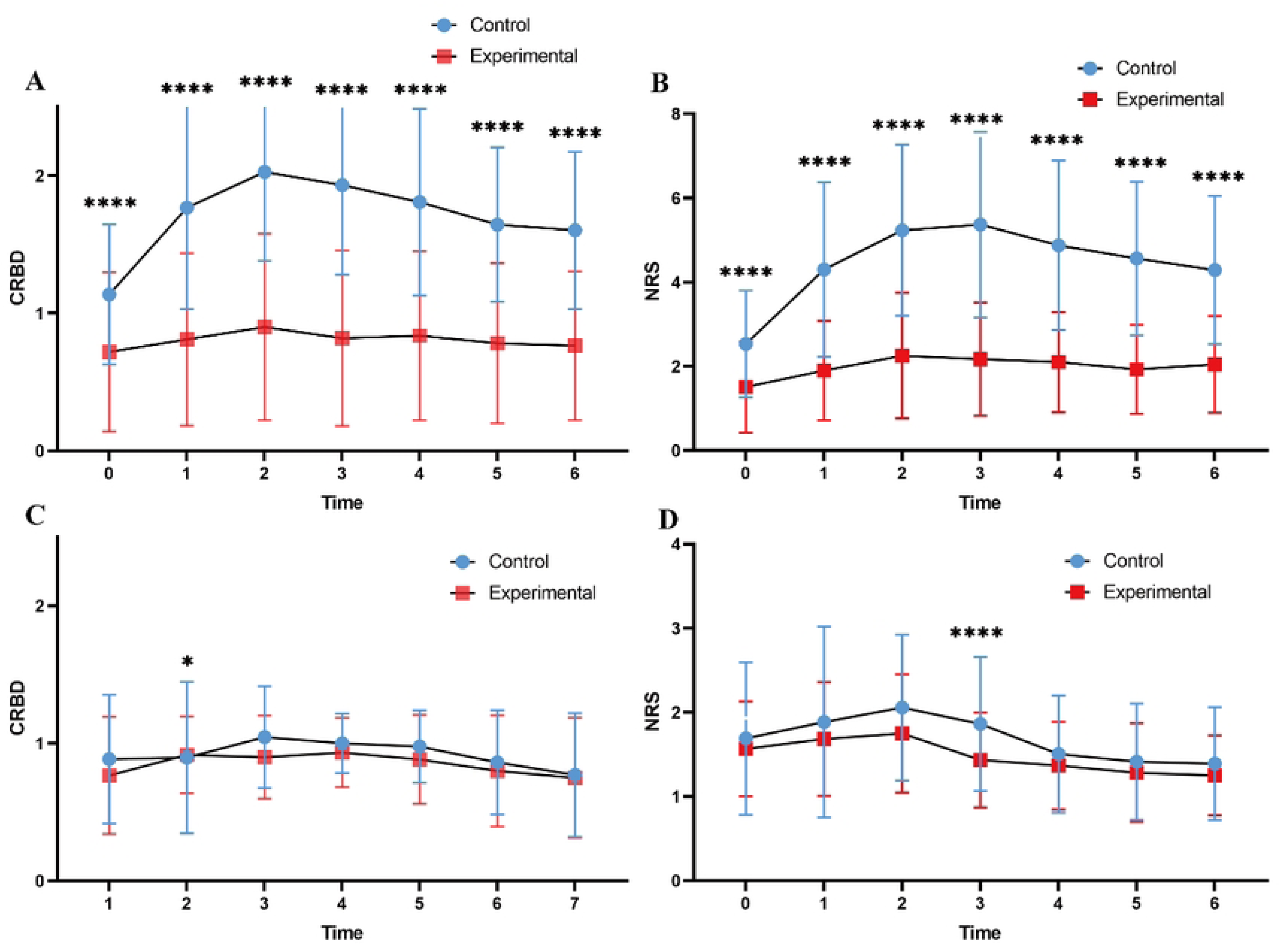
Anesthetics were effective for male patients, but not for female. Anesthetics were effective for male patients but not for female patients. Anesthetics significantly reduced CRBD (A) and NRS (B) scores in the male subgroup, but had no effect on CRBD (C) and NRS (D) scores in the female subgroup. CRBD, catheter-related bladder discomfort; NRS, numerical rating scale.

### 4. Anesthetics were effective for both ureteroscopy and flexible endoscopy

When analyzing by surgical approach, we found the use of local anesthetics significantly reduced postoperative CRBD scores and urethral pain NRS scores in both the Ureteral Rigid Ureteroscopy Lithotripsy subgroup and the Ureteral Flexible Ureteroscopy Lithotripsy subgroup (all P < 0.01, Fig. 4A-D). Without of local anesthetics, no significant difference in postoperative CRBD scores and urethral pain NRS scores observed between the two subgroups (Supplementary Table1). Similarly, when local anesthetics administered, no significant difference in postoperative CRBD scores and urethral pain NRS scores observed between the two subgroups (Supplementary Table 2). These results showed that, under the same conditions, both Ureteral Rigid Ureteroscopy Lithotripsy and Ureteral Flexible Ureteroscopy Lithotripsy have similar effects on postoperative CRBD scores and urethral pain NRS scores.

**Fig. 4.**
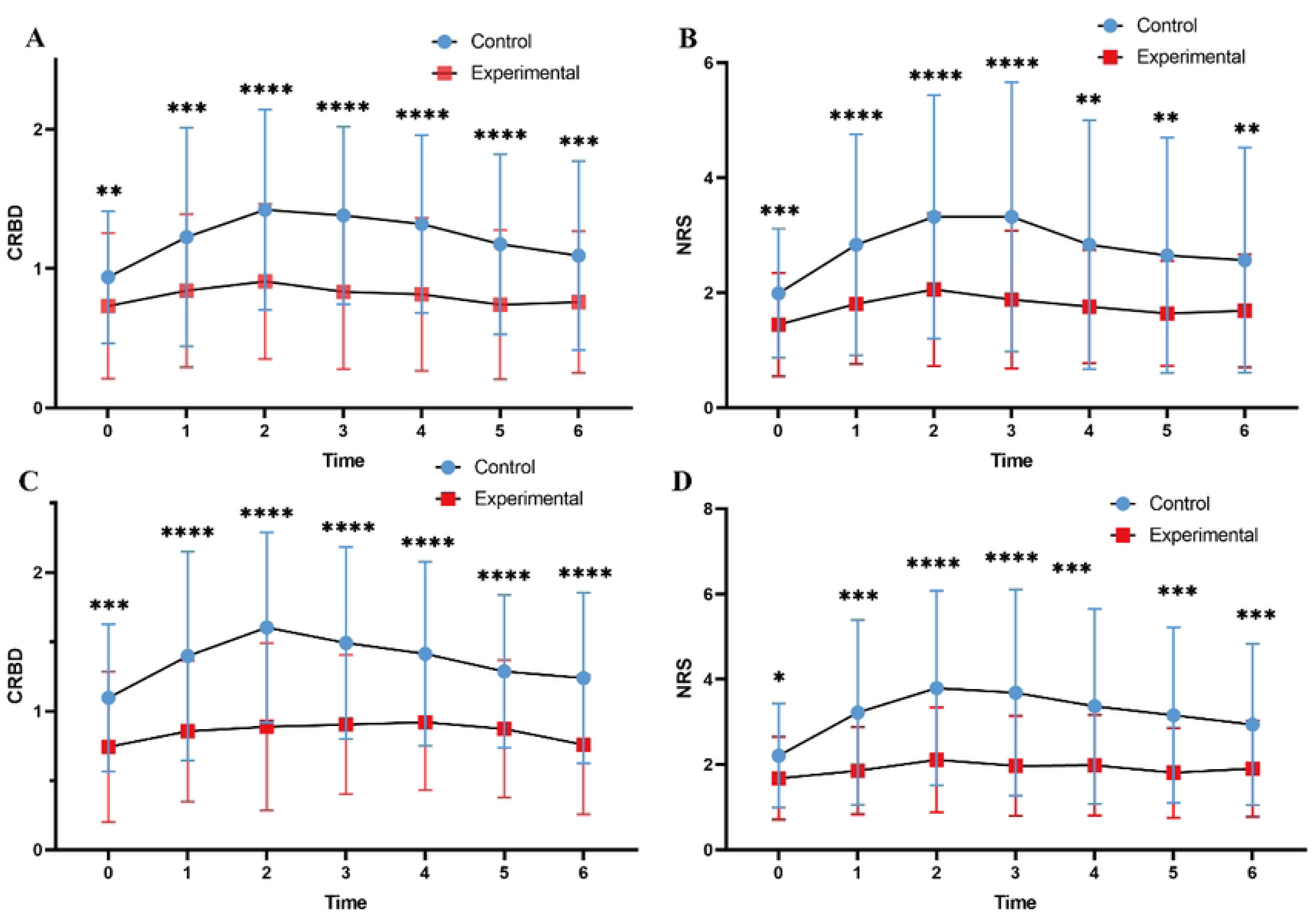
Anesthetics were effective for ureteroscopy and flexible endoscopy. Local anesthetics significantly reduced postoperative CRBD scores and urethral pain NRS scores in both the Ureteral Rigid Ureteroscopy Lithotripsy subgroup (A, B) and the Ureteral Flexible Ureteroscopy Lithotripsy (C, D) subgroup. CRBD, catheter-related bladder discomfort; NRS, numerical rating scale.

### 5. Different anesthetics exhibited no notable discrepancies

Subgroup analysis within the experimental group presented no significant difference in CRBD scores and urethral pain NRS scores between the Tetracaine Hydrochloride Gel Group and the Oxybuprocaine Gel Group (P < 0.01, Fig. 5A, B), suggesting the treatment effect was consistent across these subgroups.

**Fig. 5.**
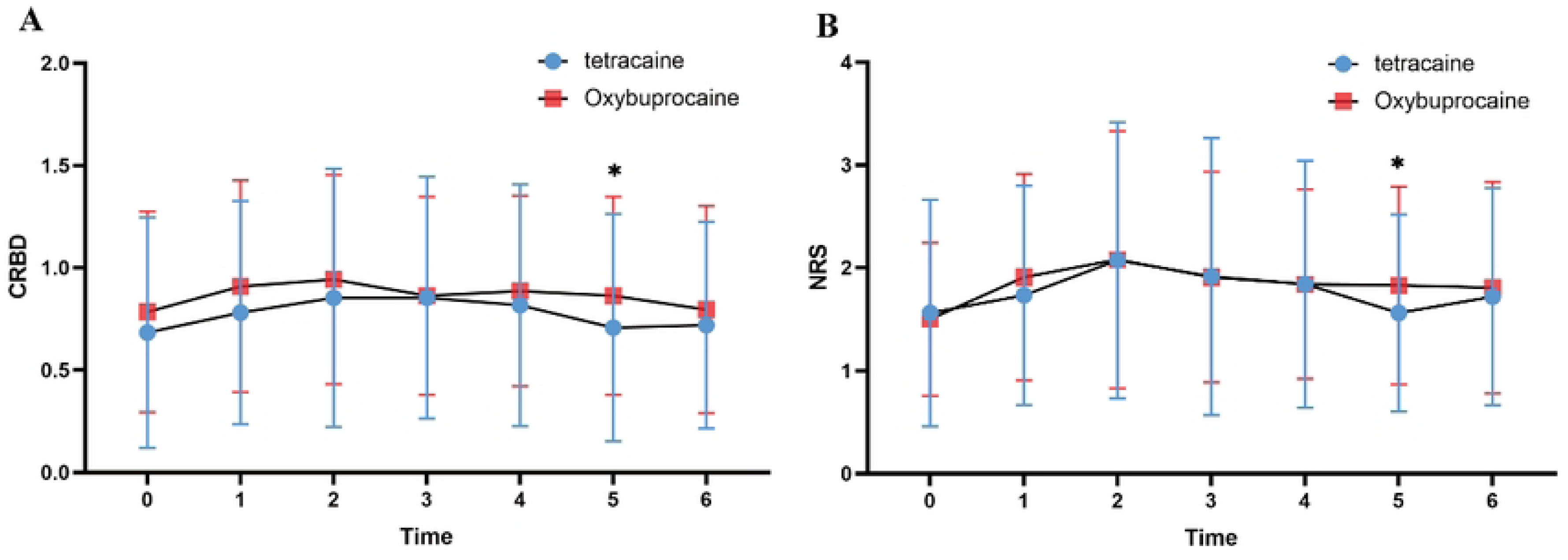
Different anesthetics exhibited no notable discrepancies. There was no significant difference in CRBD (A) and NRS (B) scores between Tetracaine Hydrochloride Gel and Oxybuprocaine Gel. CRBD, catheter-related bladder discomfort; NRS, numerical rating scale.

## Discussion

CRBD is a common complication following procedures such as transurethral surgeries, ureteral stent placement, indwelling urinary catheters, and the presence of stone fragments [1, 10, 11, 16]. These reasons contribute to bladder irritation and discomfort, leading to the symptoms of CRBD. The symptoms and underlying mechanisms of CRBD closely resemble those of OAB, characterized by urinary urgency, frequency, and bladder hypersensitivity [1, 2, 12]. Previous studies have shown that several pharmacological agents, including tolterodine, hyoscine, lidocaine, gabapentin, dexmedetomidine, diclofenac, and nefopam, are effective in relieving the symptoms of CRBD [1, 2, 5, 7, 12]. These agents target the underlying mechanisms of bladder discomfort, providing symptomatic relief for patients who undergo procedures involving catheterization or urethral stents.

Ureteroscopic lithotripsy is a common transurethral procedure used to treat upper urinary tract calculi. Following this procedure, the placement of urethral catheters and ureteral stents is essential for helping stone passage and preventing urinary tract obstruction. However, this procedure can lead to significant postoperative bladder irritation, resulting in CRBD [1, 2, 10, 12, 16]. Despite the frequent instance of CRBD after ureteroscopic lithotripsy, there is a noticeable lack of studies specifically finding out CRBD of upper urinary tract calculi management. Therefore, this study focuses on managing CRBD following ureteroscopic lithotripsy. To minimize the risk of infection and standardize study conditions, we used ureteral stents (F6) and urinary catheters (F16) from the same manufacturer. This approach ensures consistency in device-related reasons that could potentially influence the development and severity of CRBD and urethral pain. We aim to address this research gap by evaluating the effects of two local anesthetics, Tetracaine Hydrochloride Gel and Oxybuprocaine Gel, on CRBD symptoms and urethral pain. Through a controlled analysis of experimental and control groups, we seek to clarify the potential role of these drugs in mitigating CRBD post-procedure, offering new insights into treatment strategies for patients undergoing ureteroscopy lithotripsy for upper urinary tract calculi.

In this study, we used two local anesthetic drugs, Tetracaine Hydrochloride Gel and Oxybuprocaine Gel. Local anesthesia results in a loss of sensation in a specific area of the body due to suppress of excitation in nerve endings or inhibition of the conduction process in peripheral nerves. The primary mechanism of action of all local anesthetics is the reduction of the rate of depolarization by decreasing the permeability of sodium ion channels [13]. Both Tetracaine Hydrochloride Gel and Oxybuprocaine Gel have widely used in ophthalmic, otolaryngologic, and dermatologic surgeries, proving efficacy [13, 15, 17–19]. Despite their broad application across various surgical fields, limited literature exists on their use in preventing CRBD after ureteroscopic lithotripsy. This study specifically examines the impact of Tetracaine Hydrochloride Gel and Oxybuprocaine Gel on CRBD following ureteroscopic stone removal, addressing a gap in current clinical knowledge and highlighting the potential benefits of these local anesthetics in managing postoperative bladder discomfort in this context.

In this study, we found that both local anesthetics, Tetracaine Hydrochloride Gel and Oxybuprocaine Gel, significantly reduced the incidence of CRBD compared to the control group. Interestingly, no significant difference observed between the Tetracaine Hydrochloride Gel and Oxybuprocaine Gel subgroups, suggesting that both anesthetics were equally effective in relieving CRBD symptoms. When analyzing the effect of gender, we noted a more pronounced decrease in CRBD among male patients, with statistically significant differences between the experimental and control groups. In contrast, no statistically significant difference found in female patients, suggesting that gender may influence the response to local anesthetic intervention for CRBD. Besides, no significant differences found across others, including age, duration of surgery, duration of anesthesia, or surgical technique (flexible vs. rigid ureteroscopic lithotripsy). This implies the effectiveness of the local anesthetics in reducing CRBD is independent of these reasons, making the treatment strategy potentially applicable to a wide range of patient populations. Besides CRBD, the urethral pain NRS score was another important result in our study. The results for urethral pain NRS scores mirrored those observed for CRBD, with both local anesthetics showing similar decreases in urethral pain NRS scores. This suggests that both anesthetics not only reduce bladder discomfort but also contribute to overall pain relief, which is crucial for improving patient comfort during the postoperative period. The parallel effects on both CRBD and urethral pain further reinforce the potential benefits of these anesthetic agents in managing multiple aspects of post-operative recovery. This study show that Tetracaine Hydrochloride Gel and Oxybuprocaine Gel achieves significant decline in CRBD and NRS scores among male populations, potentially mediated by gender-specific urethral anatomy (notably increased urethral length and the presence of three anatomical narrowings)[2, 20]. The potential confounding effects of psychosocially-mediated conditions, particularly non-inflammatory prostatitis and related disorders, on CRBD and NRS scores measurements warrant systematic investigation through controlled longitudinal studies to decide their clinical significance.

Further, we surveyed more features, including blood pressure, heart rate, respiratory rate, and oxygen saturation. No significant differences observed between the experimental and control groups in these measures, showing the local anesthetics did not induce clinically relevant adverse effects on cardiovascular or respiratory function. We also considered the impact of comorbidities, such as hypertension and diabetes, but found no significant differences in blood pressure between the groups in hypertensive patients. Due to the small number of diabetic patients, further analysis was not conducted; however, this remains an area for future investigation.

In this study, we achieved notable findings; however, we notice several limits. The limits include its single-center design, limited sample size and Different surgeons, which may reduce the generalizability of the findings. These reasons highlight the need for further support through multicenter studies with larger sample sizes to strengthen the robustness and generalizability of the conclusions.

## Conclusions

In conclusion, to administer of Tetracaine Hydrochloride Gel or Oxybuprocaine Gel via intravesical instillation to the urethra and bladder after ureteroscopic lithotripsy significantly relieves CRBD and urethral pain, with more pronounced effects noted in male patients. Therefore, Tetracaine Hydrochloride Gel or Oxybuprocaine Gel can considered effective choices for reducing CRBD and urethral pain in patients following ureteroscopic stone removal, offering a valuable strategy for improving postoperative comfort.

## Declarations

### Ethics approval and consent to participate

The study protocol was approved by the Ethics Committee and all participants gave informed consent. (Ethics committee institution: Medical Ethics Committee of Guangxi International Zhuang Medical Hospital, Date: September 19,2024).

After being approved by the ethics committee, the research began and each participant signed an informed consent form. It was registered in the “Chinese Clinical Trial Registry” at the same time and was completed with registration on July 4, 2025(*Registration number: ChiCTR2500105477*)

## Consent for publication

Not applicable.

## Availability of data and materials

Any further questions can be directed to the corresponding authors.

## Authors’ contributions

Conceptualization: Mengxiao Wei, Fubo Wang, Wenwu Liu.

Data curation: Chenzhong Ma, Xuemei Li, Yanling Feng, Yuee Luo, Xiao Lu, Weiru Wang,

Formal analysis: Chenzhong Ma, Mengxiao Wei, Zuheng Wang, Xiao Lu, Shijia Zhou, Wenwu Liu. Funding acquisition: Fubo Wang, Wenwu Liu.

Investigation: Chenzhong Ma, Mengxiao Wei, Zuheng Wang, Xuemei Li, Yuee Luo, Weiru Wang, Shijia Zhou, Xiuping Li.

Methodology: Chenzhong Ma, Zuheng Wang, Wenwu Liu.

Project administration: Mengxiao Wei, Yanling Feng, Xiuping Li, Fubo Wang, Resources: Fubo Wang, Wenwu Liu.

Software: Chenzhong Ma, Zuheng Wang,

Supervision: Chenzhong Ma, Xuemei Li, Yanling Feng, Xiao Lu, Shijia Zhou, Xiuping Li. Validation: Mengxiao Wei, Xuemei Li, Yanling Feng, Yuee Luo, Xiao Lu, Weiru Wang, Visualization: Chenzhong Ma, Mengxiao Wei, Xuemei Li, Yuee Luo, Weiru Wang,

Writing – original draft: Chenzhong Ma, Mengxiao Wei, Zuheng Wang, Fubo Wang, Wenwu Liu. Writing – review & editing: Chenzhong Ma, Zuheng Wang, Fubo Wang, Wenwu Liu.

## Data Availability

This information will only be available after acceptance.

## Acknowledgement

We thank the patients and clinical staffs for their selfless efforts.

